# Willingness to pay for COVID-19 vaccine and its correlates: A cross-sectional survey in Bangladesh

**DOI:** 10.1101/2024.12.03.24318442

**Authors:** Mohammad Bellal Hossain, Md. Zakiul Alam, Md. Syful Islam, Shafayat Sultan, Md. Mahir Faysal, Sharmin Rima, Md. Anwer Hossain, Abdullah Al Mamun, Abdullah-Al- Mamun

## Abstract

The Government of Bangladesh has offered COVID-19 vaccines at no cost; however, sustaining this free vaccination program for a large population poses significant challenges. Assessing willingness to pay (WTP) is essential for understanding potential pricing strategies, subsidy requirements, and vaccine demand. This study aimed to estimate the prevalence of WTP for COVID-19 vaccines and identify its influencing factors to support program sustainability. Using a cross-sectional design, data were collected from 1,497 respondents through online, and face-to-face interviews and multiple logistic regression was employed to analyze the correlates of WTP. Results showed that 50.9% of participants were willing to pay, with an average WTP of 754.55 BDT (US$8.93) and a median of 300 BDT (US$3.55). WTP was significantly higher among individuals with graduate (aOR=2.2, P=0.007) or Masters & MPhil/PhD education (aOR=2, P=0.030), higher family income (aOR=1, P=0.039), and those with more excellent knowledge about the vaccine (aOR=1.1, P=0.003), positive behavioral practices (aOR=1.1, P<0.001), stronger subjective norms (aOR=1.2, P=0.009), higher anticipated regret (aOR=1.2, P=0.005), and perceived benefits (aOR=1.1, P=0.029). Conversely, WTP was lower among participants with negative attitudes toward vaccines (aOR=0.9, P<0.001) and high behavioral control (aOR=0.9, P=0.006). With nearly half of respondents unwilling to pay, the study highlights the need to improve vaccine-related knowledge, promote positive behaviors, reduce vaccine hesitancy, and enhance income-based affordability to increase WTP. Health promotion efforts should focus on disseminating vaccine knowledge and addressing negative perceptions. Additionally, a subsidized program for low-income groups could help mitigate financial barriers and promote equitable vaccine access.

## 1. Introduction

Coronavirus (COVID-19) has already caused tremendous health and socio-economic challenges worldwide. As the treatment of COVID-19 is relatively expensive, preventing COVID-19 with a safe and effective vaccine has been given the most importance (1), which is considered the most successful public health intervention (2). Even with safe and effective vaccines, the vaccination coverage program’s success relies on several other factors, such as willingness to vaccinate (3,4). The intention to be vaccinated and the success of vaccination coverage programs have been influenced by the economic considerations of many people (5,6). In this regard, willingness to pay (WTP) emerged as a concept where it is defined as the maximum amount of money an individual is willing to pay for a vaccine, health services, or technology (7–9).

To battle against the vulnerability caused by COVID-19, the Government of Bangladesh (GoB) has launched its largest-ever vaccination program, providing vaccines free of charge. However, sustaining this free vaccination program is challenging (10) for a developing country with a large population like Bangladesh. On the other hand, from the demand side perspective, individual health out-of-pocket expenditures in Bangladesh are already 74%, and 24.3% of the total population lives below the poverty line. (11,12), making fully paying for vaccines impossible. Thus, assessing WTP can offer insights into public demand and guide GoB’s future pricing and payment strategies (1,3).

Few studies in Bangladesh have measured WTP in the non-COVID situation (6,13,14), and there is much evidence of assessing WTP in the COVID-19 context in various countries (Sallam et al., 2022; Dias-Godói et al., 2021; Berghea et al., 2020; García & Cerda, 2020; Lin et al., 2020). WTP, both in COVID and non-COVID contexts, is determined by socio-economic and demographic factors (3,6,8,16,17), health-related factors, knowledge about the disease and vaccine (5), different constructs of the health belief model, and behavioral factors (9).

However, there is scanty evidence regarding WTP toward the COVID-19 vaccine in Bangladesh. Existing studies only showed the prevalence and median WTP toward the COVID-19 vaccine (18) and associated factors (19) but lacked representativeness of the Bangladeshi population as the study was only online based. Thus, our study initiated a nationwide survey to assess the prevalence of WTP regarding the COVID-19 vaccine and its associated correlates. It has important public health policy implications to identify vulnerable groups and determine a strategy to create an optimum situation where government and individual health expenditures balance.

## 2. Methods

### 2.1 Study Design and Data Collection

A cross-sectional research design was adopted in this study. The calculated sample size was 1635 using (Z^2^pq/e^2^)*Deff*NR-formula. Where Z-score for a 95% confidence interval was 1.96, the prevalence of willingness to accept a COVID-19 vaccine from an earlier study, p= 32.5% (20). We used a margin of error, e= 0.03, for sampling variation design effect (Deff) = 1.6 and a 10% non-response rate.

As GoB intended to initiate a mass vaccination program from February 7, we fixed 1 to February 7 for data collection time. Data were collected online and through face-to-face interviews through Google Forms using Bengali. The survey form link was circulated within the networks of research team members through E-mail, Facebook, WhatsApp, and other platforms. Respondents were also requested to share the link with their networks to reach maximum people. After three days, data were checked for the divisional and age-sex-specific distribution. Then, face-to-face interviews were started to determine the population’s national representation in terms of age, sex, residence, division, and marital status using quota sampling. Data were collected from randomly selected two districts of each of the eight divisions. Four days were allocated to collect data from face-to-face interviews, and interviewers maintained proper health measures while conducting interviews.

For offline data collection, the selection criteria to participate in this study were to be at least 18 years old and know about the COVID—19 vaccine—another additional criterion for the online respondents’ reading and writing ability. During the interview, 112 respondents did not consent, and another 26 did not know about the COVID-19 vaccine. Therefore, the final sample size was 1497.

### 2.2 Measures

#### 2.2.1 Outcome Variables

Two questions were used to assess the respondents’ WTP. The first question was, “Would you like to pay for the COVID-19 vaccine?” with a binary response (Yes/ No). If the response to the first question was ‘yes,’ then the second question was “What is the maximum amount you are willing to pay for the COVID-19 vaccine?”.

#### 2.2.2 Independent Variables

##### Socio-economic and demographic variables

Several socio-economic and demographic variables, such as age, sex, religion, marital status, place of residence, household income, and occupation, were used as independent variables in this study.

##### Health-related variables

Respondents’ perceived health status (3 categories) and infection status from COVID-19 of self, family members, and friends (3 binary questions) were used as respondents’ health-related independent variables.

##### Knowledge of COVID-19 vaccine

Knowledge of the COVID-19 vaccine was assessed using four Likert-type items. The scores ranged between 1 and 20, with a higher score indicating greater knowledge. The Cronbach alpha (α) was 0.643.

##### Knowledge about the Vaccination Process

Six binary (yes=1, no=0) questions were used to measure knowledge about the COVID-19 vaccination process where Cronbach alpha (α) was 0.765 with good internal consistency. The higher the scores indicate a better understanding.

##### COVID-19 Vaccine Conspiracy

Nine Likert-type items were used to assess conspiracy related to the COVID-19 vaccine (α=0.716), ranging between 9 and 45 scores, where a higher score indicates higher conspiracy toward the COVID-19 vaccine.

##### Preventive behavioral practices related to COVID-19

Preventive behavioral practices related to COVID-19 were measured using three items, ranging between 1 and 12. A higher score indicated better preventive practices with the Cronbach alpha (α=0.857).

##### Theory of Planned Behavior

*Attitude towards COVID-19 Vaccine* Six Likert-type items were used to assess COVID-19 vaccine-related attitudes (α=0.739), ranging between 6 and 30 scores. A higher score on this scale directs higher negative attitudes toward the COVID-19 vaccine.

*Subjective Norm* Subjective norm toward the COVID-19 vaccine was assessed using one 5-point item: I believe my family members will support me in getting vaccinated against COVID-19.

*Perceived Behavioral Control was measured using one 5-point item.I can register for COVID-19 vaccination if I want*.

*Anticipated Regret* The regret of not getting vaccinated was assessed using one 5-point item: If I do not get a COVID-19 vaccine and end up getting Coronavirus, I will regret not getting the vaccination.

##### Health Belief Model

The Health Belief Model was constructed by four components: Perceived Susceptibility, Perceived Severity, Perceived Benefits, and Perceived Barriers.

*Perceived Susceptibility* Perceived Susceptibility of COVID-19 was measured by two 5-point Likert scale questions where Cronbach alpha, α is 0.657.

*Perceived Severity:* Two 5-point Likert-type items were used to measure the perceived severity of COVID-19 where Cronbach alpha, α is 0.612.

*Perceived Benefits:* Three 5-point Likert scale questions were used to measure perceived benefits of the COVID-19 vaccination where Cronbach alpha, α is 0.841.

*Perceived Barriers:* Five 5-point Likert scale questions were used to measure the perceived barriers (α=0.739) to getting the COVID-19 vaccination.

### 2.3 Statistical Analysis

We first employed univariate descriptive statistics (Percentage, mean, standard deviation-SD) for all the variables. At the bivariate level for WTP, the Chi-square test and point Bi-serial Correlation were employed, and then statistically significant variables (P≤0.05) at the bivariate level were entered into a hierarchical logistic regression model to assess WTP determinants. We used only descriptive analysis (mean and Median) for WTP’s highest amount of money. Data were analyzed using SPSS, version 26.

### 2.4 Ethical Approval

Ethical approval was taken from the Bangladesh Medical Research Council-BMRC (Registration No. 39131012021). There was voluntary participation in the research, and no incentives were provided to the participants. Firstly, the aims, objectives, potential scopes, and implications of the findings of this study were communicated to the participants, and then the participants provided their written consent. After that, the respondents participated in the study.

## 3. Results

### 3.1 Background characteristics of the participants

The mean age of the respondents was 33.67 (SD=12.94), where more than half of the respondents (53.8%) were male (Table 1). About 86.9% of the respondents were Muslim, and nearly two-thirds (61.6%) were married. Most respondents (79.2%) had at least secondary and higher secondary education. Almost two-thirds (64.3%) of respondents were from rural areas, and the highest percentage of respondents (31.9%) were from the Dhaka division (Table 1). One-third (31.6%) of the respondents were students and unemployed. The respondents’ mean number of household members was 4.95, and they had a mean collective monthly family income of 37,627 BDT. About 58.7% of the respondents identified themselves as having good health status. The mean score of knowledge about the COVID-19 vaccine, vaccine process, vaccine conspiracy, and behavioral practice were 11.40, 2.84, 12.65, and 8.80, respectively (Table 1).

**Table 1:** Background characteristics, prevalence and differentials of WTP for COVID-19 vaccine among the study population (n= 1497).

| Variables | Study sample<br>N (%) / mean (SD) | Willingness To Pay |  | p-value |
| --- | --- | --- | --- | --- |
|  |  | No (%) | Yes (%) |  |
| **Sex** |  |  |  | 0.717 |
| Female | 692 (46.2) | 329 (47.5) | 363 (52.5) |  |
| Male | 805 (53.8) | 391 (48.6) | 414 (51.4) |  |
| **Religion** |  |  |  | <0.001 |
| Muslim | 1301 (86.9) | 650 (50) | 651 (50) |  |
| Others | 196 (13.1) | 70 (35.7) | 126 (64.3) |  |
| **Marital status** |  |  |  | 0.009 |
| Unmarried | 575 (38.4) | 252 (43.8) | 323 (56.2) |  |
| Married | 922 (61.6) | 468 (50.8) | 454 (49.2) |  |
| **Education** |  |  |  | <0.001 |
| No education | 129 (8.6) | 90 (69.8) | 39 (30.2) |  |
| Primary | 179 (12.0) | 112 (62.6) | 67 (37.4) |  |
| Secondary and higher secondary | 448 (29.9) | 236 (52.70) | 212 (47.30) |  |
| Graduate | 400 (26.7) | 156 (39) | 244 (61) |  |
| Masters and MPhil/PhD | 341 (22.8) | 126 (37) | 215 (63) |  |
| **Place of residence** |  |  |  | <0.001 |
| Rural | 963 (64.3) | 510 (53) | 453 (47) |  |
| Urban (other than city corporation) | 179 (12) | 66 (36.9) | 113 (63.1) |  |
| City Corporation | 355 (23.7) | 144 (40.6%) | 211 (59.4) |  |
| **Administrative division** |  |  |  | 0.004 |
| Barisal | 114 (7.6) | 67 (58.8) | 47 (41.2) |  |
| Chattogram | 253 (16.9) | 114 (45.1) | 139 (54.9) |  |
| Dhaka | 478 (31.9) | 209 (43.7) | 269 (56.3) |  |
| Khulna | 137 (9.2) | 75 (54.7) | 62 (45.3) |  |
| Mymensingh | 108 (7.2) | 64 (59.3) | 44 (40.7) |  |
| Rajshahi | 180 (12.0) | 88 (48.9) | 92 (51.1) |  |
| Rangpur | 114 (7.6) | 58 (50.9) | 56 (49.1) |  |
| Sylhet | 113 (7.5) | 45 (39.8) | 68 (60.2) |  |
| **Occupation** |  |  |  | <0.001 |
| Government, private, & NGO sector job | 202 (13.5) | 81 (40.1) | 121 (59.9) |  |
| Professional | 277 (18.5) | 115 (41.5) | 162 (58.5) |  |
| Homemakers | 348 (23.2) | 195 (56) | 153 (44) |  |
| Students and unemployed | 473 (31.6) | 210 (44.4) | 263 (55.6) |  |
| Agriculture, Day Laborer | 102 (6.81) | 65 (63.7) | 37 (36.3) |  |
| Others | 95 (6.34) | 54 (56.8) | 41 (43.2) |  |
| **Perceived health status** |  |  |  | 0.073 |
| Bad/ very bad | 51 (3.4) | 27 (52.9) | 24 (47.1) |  |
| Moderate | 333 (22.2) | 177 (53.2) | 156 (46.8) |  |
| Good/ very good | 1113 (74.3) | 516 (46.4) | 597 (53.6) |  |
| **Was infected with the Coronavirus** |  |  |  | 0.026 |
| Yes | 86 (5.7) | 31 (36.0) | 55 (64.0) |  |
| No | 1411 (94.3) | 689 (48.8) | 722 (51.2) |  |
| **Respondent's family members were infected with Coronavirus** |  |  |  | 0.016 |
| Yes | 152 (10.2) | 59 (38.8) | 93 (61.2) |  |
| No | 1345 (89.8) | 661 (49.1) | 684 (50.9) |  |
| **Respondent's friends got infected with Coronavirus** |  |  |  | <0.001 |
| Yes | 600 (40.1) | 240 (40) | 360 (60) |  |
| No | 897 (59.9) | 480 (53.5) | 417 (46.5) |  |
| **Age (r)** | 33.67 (12.94) | -.060 |  | 0.020 |
| **Household size (r)** | 4.95 (1.98) | -0.030 |  | 0.242 |
| **Monthly household income (r)** | 37627 (81296) | .123 |  | <0.001 |
| **Knowledge about the COVID-19 vaccine** | 11.40 (2.19) | .093 |  | <0.001 |
| **Knowledge about the vaccination process** | 2.84 (1.97) | .195 |  | <0.001 |
| **Covid-19 vaccine conspiracy** | 12.65 (3.69) | -.148 |  | <0.001 |
| **Behavioral practice to prevent COVID-19** | 8.80 (2.71) | .230 |  | <0.001 |
| **Total** | **1497 (100)** | **720 (48.1)** | **777 (50.9)** |  |

### 3.2 The prevalence of willingness to pay (WTP) for the COVID-19 Vaccine

The prevalence of willingness to pay for the COVID-19 vaccine was 50.9%. However, almost half of the respondents (48.1%) refused to pay any money for the COVID-19 vaccine.

### 3.3 Differentials of WTP for COVID-19 vaccine

WTP was statistically significantly (P ≤ 0.05) varied by age, religion, marital status, education, place of residence, administrative division, occupation, coronavirus infection status of respondent, family & friends, household size, and household income (Table 1). Knowledge about the COVID-19 vaccine and vaccination process, COVID-19 conspiracy beliefs, behavioral practice, attitude toward a vaccine, subjective norm, perceived behavioral control, anticipated regret, perceived susceptibility, severity, benefits, and barriers were statistically significantly correlated with WTP.

### 3.4 Correlates of WTP for COVID-19 vaccine

The significant variables at the bivariate level were entered into a hierarchical logistic regression model to assess correlates of WTP for the COVID-19 vaccine. We produced three models in this study. The first model was constructed with variables such as socio-economic and demographic, knowledge of COVID-19 vaccine and vaccination process, COVID-19 Vaccine Conspiracy, and preventive behavioral practices related to COVID-19. The second model was constructed with the variables of the first model and components of the theory of planned behavior. Furthermore, the final model included all second and health belief model variables (perceived susceptibility, severity, barrier, benefits, and health-related variables). The final model showed that religion, education, administrative division of Bangladesh, household income, knowledge about the COVID-19 vaccine & vaccination process, behavioral practice, attitude toward a vaccine, subjective norms, perceived behavioral control, anticipated regret, and barriers were statistically significant (P ≤ 0.05) predictors of WTP (Table 2).

**Table 2:** Correlates of willingness to pay (WTP) for COVID-19 vaccine in Bangladesh.

| Variables | Model 1<br>aOR | Model 2<br>aOR | Model 3<br>aOR |
| --- | --- | --- | --- |
| Age | 1.0 (1.0, 1.0) | 1.0 (1.0, 1.0) | 1.0 (1.0, 1.0) |
| Religion other (Muslim as RC) | 1.8 (1.3, 2.6)** | 1.5 (1.1, 2.2)* | 1.5 (1.0, 2.1)* |
| Marital status: unmarried (married as RC) | 1.0 (0.7, 1.5) | 1.1 (0.8, 1.7) | 1.1 (0.8, 1.7) |
| Education (no education as RC) |  |  |  |
| Primary | 1.2 (0.7, 1.9) | 1.1 (0.6, 1.8) | 1.0 (0.6, 1.7) |
| Secondary and higher secondary | 1.5 (0.9, 2.5) | 1.3 (0.8, 2.3) | 1.3 (0.8, 2.2) |
| Graduate | 2.4 (1.4, 4.1)** | 2.3 (1.3, 4.1)** | 2.2 (1.2, 4.0)** |
| Masters & MPhil/ PhD | 2.0 (1.1, 3.6)* | 2.0 (1.1, 3.7)* | 2.0 (1.1, 3.6)* |
| **Place of residence (Rural as RC)** |  |  |  |
| Urban (other than CC) | 1.2 (0.8, 1.7) | 1.2 (0.8, 1.7) | 1.1 (0.8, 1.7) |
| City Corporation | 1.0 (0.7, 1.4) | 1.1 (0.8, 1.6) | 1.1 (0.8, 1.6) |
| **Administrative division of Bangladesh (Barisal as RC)** |  |  |  |
| Chattogram | 2.3 (1.4, 3.8)** | 2.5 (1.5, 4.1)** | 2.5 (1.5, 4.3)** |
| Dhaka | 2.1 (1.3, 3.3)** | 2.3 (1.4, 3.7)** | 2.3 (1.4, 3.8)** |
| Khulna | 1.8 (1.1, 3.2)* | 2.4 (1.4, 4.4)** | 2.5 (1.4, 4.5)** |
| Mymensingh | 1.0 (0.6, 1.9) | 1.1 (0.6, 2.0) | 1.1 (0.6, 2.1) |
| Rajshahi | 2.3 (1.3, 3.8)** | 2.6 (1.5, 4.6)** | 2.7 (1.5, 4.7)** |
| Rangpur | 1.9 (1.0, 3.3)* | 2.4 (1.3, 4.4)** | 2.4 (1.3, 4.6)** |
| Sylhet | 3.3 (1.9, 6.0)*** | 4.0 (2.2, 7.4)*** | 4.1 (2.2, 7.7)*** |
| **Occupation ( government, private, NGO sector job as RC)** |  |  |  |
| Professional | 1.1 (0.8, 1.7) | 1.1 (0.7, 1.7) | 1.1 (0.7, 1.7) |
| Homemakers | 1.4 (0.9, 2.3) | 1.4 (0.9, 2.3) | 1.5 (0.9, 2.4) |
| Students & unemployed | 1.1 (0.7, 1.7) | 1.0 (0.7, 1.7) | 1.1 (0.7, 1.8) |
| Agriculture and day labor | 1.2 (0.7, 2.2) | 1.2 (0.6, 2.2) | 1.1 (0.6, 2.1) |
| Others | 0.9 (0.5, 1.6) | 0.9 (0.5, 1.5) | 0.8 (0.5, 1.5) |
| **Income** | 1.0 (1.0, 1.0)* | 1.0 (1.0, 1.0)* | 1.0 (1.0, 1.0)* |
| **Knowledge about the COVID-19 vaccine** | 1.1 (1.1, 1.2)*** | 1.1 (1.0, 1.2)** | 1.1 (1.0, 1.2)** |
| **Knowledge about the vaccination process** | 1.1 (1.1, 1.2)*** | 1.1 (1.0, 1.2)* | 1.1 (1.0, 1.2) |
| **Behavioral practice to prevent COVID-19** | 1.1 (1.1, 1.2)*** | 1.1 (1.0, 1.2)*** | 1.1 (1.0, 1.2)*** |
| **Covid-19 vaccine conspiracy** | 0.9 (0.9, 1.0)*** | 1.0 (1.0, 1.0) | 1.0 (1.0, 1.0) |
| **Attitude toward vaccine** |  | 0.9 (0.9, 0.9)*** | 0.9 (0.9, 0.9)*** |
| **Subjective norms** |  | 1.3 (1.1, 1.4)** | 1.2 (1.1, 1.4)** |
| **Perceived behavioral control** |  | 0.8 (0.8, 0.9)** | 0.9 (0.8, 1.0)** |
| **Anticipated regret** |  | 1.2 (1.1, 1.3)** | 1.2 (1.1, 1.3)** |
| **Susceptibility** |  |  | 0.9 (0.9, 1.0) |
| **Severity** |  |  | 1.0 (0.9, 1.1) |
| **Benefits** |  |  | 1.1 (1.0, 1.1)* |
| **Barriers** |  |  | 1.0 (0.9, 1.0) |
| **Respondent got infected with Coronavirus ( no as RC)** |  |  | 0.9 (0.5, 1.5) |
| **Respondent's family member got infected with Coronavirus (no as RC)** |  |  | 0.9 (0.6, 1.4) |
| **Respondents friends or peers got infected with Coronavirus (no as RC)** |  |  | 0.8 (0.6, 1.1) |
| Constant | 0.034*** | 0.042*** | 0.044*** |
| **Model Summary** |  |  |  |
| N | 1497 | 1497 | 1497 |
| -2 Log likelihood | 1851 | 1753 | 1744 |
| Cox & Snell R Square | 0.138 | 0.192 | 0.197 |
| Nagelkerke R Square | 0.184 | 0.257 | 0.263 |

aOR= Adjusted Odds Ratio, RC= Reference category, \*= p<0.05, \*\*= p<0.01, \*\*\*= p<0.001

According to the final model, respondents of other religions were 50% more likely to pay for the COVID-19 vaccine than Muslims (P= 0.037). Similarly, education was a statistically significant (P= 0.008) predictor of WTP, where respondents who had Graduate (aOR= 2.2, P= 0.007), Masters & MPhil/PhD (aOR= 2.0, P= 0.030) degrees had higher WTP than respondents who had no education (Table 2). Regional variation of WTP was found and division was a significant predictor of WTP (P<0.001), which showed respondents from Chattogram (aOR=2.5, P=0.001), Dhaka (aOR=2.3, P=0.001), Khulna (aOR=2.5, P=0.003), Rajshahi (aOR=2.7, P=0.001), Rangpur (aOR=2.4, P=0.005) and Sylhet (aOR= 4.1, P<0.001) had higher WTP than respondents from Barisal (Table 2). Our results showed that WTP increased with household income (aOR=1.0, P=0.039). Findings showed that with increasing level of knowledge about the COVID-19 vaccine (P=0.003), Behavioral preventive practices (P<0.001), higher subjective norm (P=0.009), higher anticipated regret (P=0.005) and increasing perceived benefit from the vaccine (P=0.029) WTP increased by 10%, 10%, 20%, and 10% respectively (Table 2). On the other hand, a higher negative attitude toward the COVID-19 vaccine (P<0.001) and higher difficulty in registration (P=0.006) decreased WTP by 10% and 10%, respectively. (Table 2).

### 3.5 Willingness to pay the highest amount of money

The mean and median willingness to pay the highest amount of money among the respondents was 754.55 BDT (US$8.93) and 300 BDT (US$3.55) for the COVID-19 vaccine. Nearly half of the respondents (48.1%) were unwilling to pay for vaccines. The willingness to pay the highest amount of money ranged from the lowest of 1 BDT (US$0.012) to the highest of 30,000 BDT (US$353.73).

Figure 1 explains explains that mean WTP by monthly household income. There was an increasing trend of WTP with increasing income except for the lowest income category (0-5000 BDT). Respondents from the 5001-10000 BDT income group had a mean WTP of 376 BDT (US $4.44). On the other hand, the mean WTP of the 50000+ BDT income group was 1479 BDT (US $17.45).

**Figure 1:**
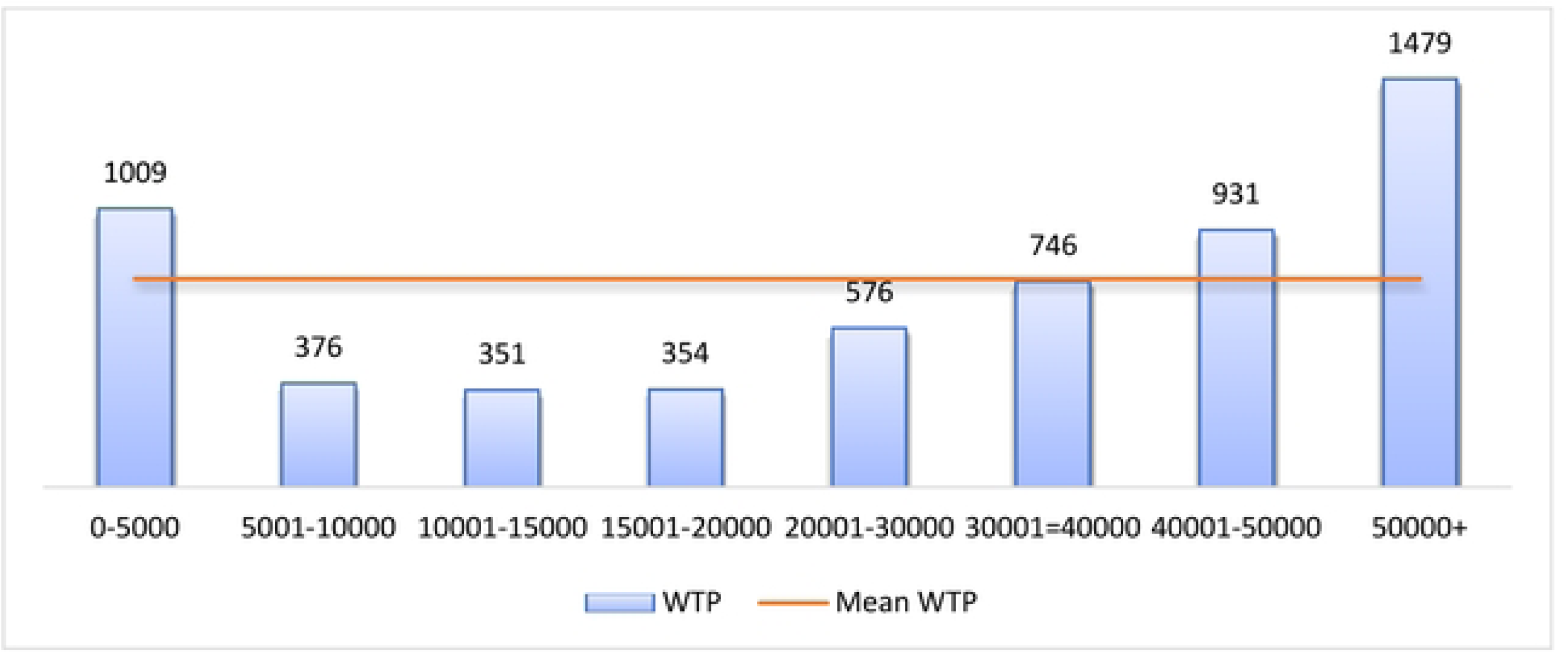
Mean WTP highest amount of money by household income category.

## 4. Discussion

A successful vaccination program depends not only on safe and effective vaccines but also on vaccine hesitancy, willingness to pay etc. Our study objective was to explore the WTP toward the COVID-19 vaccine and its associated factors to identify the vulnerable groups and possible interventions. In this regard, Our study demonstrated that half of the respondents (50.9 %) willingly paid for the COVID-19 vaccine in Bangladesh. The mean and median WTP were 754.55 BDT (US$ 8.93) and 300 BDT (US$ 3.55). However, an existing online survey on WTP found a greater prevalence of WTP (68.4%) toward the COVID-19 vaccine than our study, and even the median WTP (U.S. $ 7.08) was also found to be higher than our findings (18). These differences might be attributed to the variation in the study population selection of the two studies. The existing study collected data through an online platform, thus leading to selection bias of the study population, but our study collected data from online and face-to-face interviews. Several studies were also found in the world context with higher prevalence and higher mean and Median of WTP than our study (3,8,15). As there is a concern about vaccine resistance and budgetary limitation, which creates an expectation that the government would fully subsidize the vaccine, this kind of expectation contributes to the lower willingness to accept and pay for the vaccine (21). However, the lower prevalence of WTP is a concerning issue to attain herd immunity because there is an estimated that perhaps 60% of the population needs to be under an immunization system to ensure the effectiveness of any vaccine (3). Our study findings found that religion significantly predicts WTP for the COVID-19 vaccine in Bangladesh. Muslims had significantly lower WTP than other religions (Hindu, Christian, and Buddhism), and the existing literature supports the findings (3). The Muslim community is known for its fatalistic view about health and appreciation, acceptance & patience for their current situation (22,23). In Bangladesh, the majority of the population is Muslim. Muslims have various religion-related conspiracy beliefs, lower perception and level of knowledge regarding health and vaccines, and trust issues on the vaccines of other non-Muslim countries, which may influence their decision to purchase a vaccine and ultimately interplay for lower WTP toward COVID-19 vaccine (24–27).

Our study shows that education significantly predicts WTP for the COVID-19 vaccine (16,17). In our study, respondents who had Graduate and Masters & MPhil/PhD level education had higher WTP than respondents who had no education. Educated people are more likely to be conscious about health and more willing to prevent diseases through preventive care such as vaccination(28–30). Education also determines the knowledge of health & vaccines, and those with a low level of knowledge cannot grow willingness to pay for vaccines (29,31). Educated people are more likely to have higher incomes, giving them higher purchasing power.

In our study, household income was positively associated with WTP for the COVID-19 vaccine in Bangladesh. Income has proved to be the most influencing factor for determining WTP both in COVID and non-COVID vaccines (Sallam et al., 2022; Harapan et al., 2020; Sarker et al., 2020; Rajamoorthy et al., 2019). Those from higher income groups have the higher purchasing power to invest more in health and practice healthier behaviors such as WTP toward vaccines(6,30). Similarly, wealthier families want to pay more to the health sector to avoid the fatality of diseases and are more interested in improving their immune system through vaccination (32). Again, higher-income individuals tend to be educated, making them conscious about health.

The study findings revealed a divisional disparity regarding willingness to pay (WTP) for the COVID-19 vaccine in Bangladesh. There is a lack of evidence of regional variation with WTP for vaccines. However, there is evidence of socio-economic and demographic contextual variation among divisions in Bangladesh (33). Barisal and Rangpur are the two divisions with the highest poverty headcount ratio, whereas Sylhet, Dhaka, Khulna, and Chattogram have much lower poverty headcount ratios (11). Comparative poor households in Barisal and Rangpur may lead to lower willingness to pay (WTP) for the COVID-19 vaccine.

Apart from the socio-economic and demographic status, the knowledge about the COVID-19 vaccine and preventive behavioral practices were associated with higher WTP, and the literature supports the present study’s findings (5,34). Increased knowledge about the COVID-19 vaccine indicates more reliable knowledge regarding vaccine’s safety and effectiveness, which increases trust in the vaccine; thus, WTP may increase. Again, people with higher preventive behavioral practices tend to be educated and health conscious, so they are expected to have higher WTP towards any vaccine (35).

The study findings show that the Theory of Planned behavior components were statistically significant predictors of WTP. The main argument of TPB is that behavior intention is the most important determinant of health behavior, in this case, WTP (36). Our study findings found that respondents with a higher negative attitude toward vaccines also had lower WTP. Attitude acts as a personal evaluation of a behavior; as a result, the intention to WTP decreases with a higher negative attitude. Similarly, perceived behavioral control indicates that performing health behavior is not solely on the respondent’s hand; in this case, it is difficult to register. In our study, difficulty in online registration acts as a structural barrier that reduces WTP for the COVID-19 vaccine. On the other hand, subjective norms and anticipated regret in our study findings positively associated with WTP for the vaccine. In our study, respondents whose family members approved their decision to take the vaccine and highly regretted getting Coronavirus disease had higher WTP, and it is also a part of subjective norms because it acts as permission from family members or friends toward a health behavior(36). Thus, intention toward a health behavior regarding a disease or vaccine is driven by attitude toward the disease or vaccine, subjective norms, perceived behavioral control etc. (36,37).

Our study found Perceived benefits from vaccines as a significant predictor of WTP. When people perceive that the disease will be responsible for high vulnerability regarding their health and economic condition and getting a vaccine can eliminate that cost (the cost of the disease, both health and economic, is higher), they are more willing to pay (WTP) for the vaccine and helps to increase WTP (36).

Similarly, our findings revealed that those who are experiencing severe morbid conditions are more likely to pay a high amount of money for the COVID-19 vaccine. The severity of the disease is associated with a higher intention to pay because those who are already going through the morbid situation are not confident about their immune system to protect from disease and are more likely to pay for an effective vaccine (7,38).

This study intends to explore the prevalence and associated factors of WTP for a COVID-19 vaccine in Bangladesh, which will help GoB and policymakers promote a successful vaccination program at a larger scale among the mass population by eliminating economic challenges. However, some limitations should be taken into account before accepting our findings. Our study adopted a cross-sectional study design, which is limited to properly generating causal inferences due to temporal issues. Again, we used non-probability sampling to reach our study population. We collected self-reported data on health status and other socio-demographic variables, which may suffer from recall bias. Finally, our study is almost nationally representative regarding age, sex, religion, residence, and marital status in Bangladesh, but it could not be represented in terms of education, occupation, or income status.

## 5. Conclusion

The study findings propose that health promotion materials and awareness campaigns as part of the Behavior Change Communication (BCC) program should be developed to increase knowledge about the COVID-19 vaccine. It will also increase preventive behavioral practices and reduce negative attitudes towards vaccines and vaccine-related conspiracy beliefs. Here, mass media can be an effective platform to circulate the accurate message of the COVID-19 vaccine, and religious leaders can also be incorporated to mitigate religion-related mistrust and misconception. Policymakers should rethink the online registration procedure for vaccine uptake because it is a structural barrier to WTP for the COVID-19 vaccine. Regarding our findings, an easy alternative system should be introduced to the mass population to achieve sustainability of the vaccination program. Authors suggest that policymakers consider a subsidization program that considers socio-economic stratification and where lower-income groups should be highlighted to reduce the catastrophic income challenge regarding WTP for the COVID-19 vaccine. Otherwise, a reasonable price for our findings should be fixed so that the COVID-19 vaccine can be affordable for all. This helps achieve the highest vaccine coverage and run a successful vaccination program without economic hardship.

## Author Contributions

### Conceptualization

Mohammad Bellal Hossain, Md. Zakiul Alam, Md. Syful Islam, Shafayat Sultan, Md. Mahir Faysal, Sharmin Rima, Md. Anwer Hossain, Abdullah Al Mamun, Abdullah-Al-Mamun.

### Data curation

Mohammad Bellal Hossain, Md. Zakiul Alam, Shafayat Sultan, Md. Mahir Faysal, Sharmin Rima, Md. Anwer Hossain, Abdullah Al Mamun.

### Formal analysis

Mohammad Bellal Hossain, Md. Zakiul Alam, Shafayat Sultan.

Investigation: Mohammad Bellal Hossain, Md. Zakiul Alam, Md. Syful Islam, Shafayat Sultan, Md. Mahir Faysal, Sharmin Rima, Md. Anwer Hossain, Abdullah Al Mamun.

### Methodology

Mohammad Bellal Hossain, Md. Zakiul Alam, Md. Syful Islam, Shafayat Sultan, Md. Mahir Faysal, Sharmin Rima, Md. Anwer Hossain, Abdullah Al Mamun, Abdullah-Al-Mamun.

### Project administration

Mohammad Bellal Hossain, Md. Zakiul Alam, Md. Syful Islam, Shafayat Sultan, Md. Mahir Faysal, Sharmin Rima, Md. Anwer Hossain, Abdullah Al Mamun.

### Resources

Mohammad Bellal Hossain, Md. Zakiul Alam, Md. Syful Islam, Shafayat Sultan, Md. Mahir Faysal, Sharmin Rima, Md. Anwer Hossain, Abdullah Al Mamun.

### Software

Md. Zakiul Alam, Abdullah Al Mamun

### Supervision

Mohammad Bellal Hossain, Md. Zakiul Alam, Md. Syful Islam, Shafayat Sultan, Md. Mahir Faysal, Sharmin Rima, Md. Anwer Hossain, Abdullah Al Mamun.

### Validation

Mohammad Bellal Hossain, Md. Zakiul Alam, Shafayat Sultan.

### Visualization

Mohammad Bellal Hossain, Md. Zakiul Alam, Shafayat Sultan.

### Writing – original draft

Mohammad Bellal Hossain, Abdullah Al Mamun, Sharmin Rima.

### Writing – review & editing

Mohammad Bellal Hossain, Md. Zakiul Alam, Md. Syful Islam, Shafayat Sultan, Md. Mahir Faysal, Sharmin Rima, Md. Anwer Hossain, Abdullah Al Mamun, Abdullah-Al-Mamun

### Institutional Review Board Statement

The study was conducted following the Declaration of Helsinki and approved by the Bangladesh Medical Research Council-BMRC (Registration No. 39131012021).

### Informed Consent Statement

Written informed consent was obtained from all the study participants.

### Data Availability Statement

The authors will make the raw data supporting this article’s conclusions available upon request.

## Acknowledgment

We would like to thank the participants in this study and the data collectors for their contributions during the challenging COVID-19 pandemic.

